# Leptin and LH/FSH Ratio as Independent Predictors of Polycystic Ovary Syndrome in Normoglycemic Women: A Case-Control Study

**DOI:** 10.1101/2025.09.11.25335551

**Authors:** Jyoti Singh, Anju Jain

## Abstract

**BACKGROUND:** Polycystic ovary syndrome (PCOS) is a multifaceted disorder characterized by reproductive, metabolic, and endocrine disturbances. While insulin resistance (IR) is recognized as a central feature, the interplay of adipokines, lipid metabolism, gonadotropin imbalance, and inflammation in normoglycemic PCOS women remains underexplored.

**METHODS:** A case-control study was conducted on 30 newly diagnosed normoglycemic women with PCOS (Rotterdam criteria, 2003) and 30 age-matched healthy controls. Anthropometric, hormonal (LH, FSH, LH/FSH ratio, prolactin, estrogen, progesterone, testosterone), metabolic (fasting glucose, HbA1c, insulin, HOMA-IR), lipid (cholesterol, triglycerides, HDL), adipokine (leptin), and hematological parameters were assessed. Group comparisons were performed using independent t-test/Mann-Whitney test as appropriate. Correlation analysis explored inter-relationships among variables, and multivariate logistic regression identified independent predictors of PCOS.

**RESULTS:** Compared with controls, PCOS women had significantly higher BMI (26.3 +/- 5.3 vs. 23.4 +/- 3.2 kg/m2; p=0.014), WHR (0.861 +/- 0.061 vs. 0.799 +/- 0.060; p<0.001), LH (7.5 +/- 3.7 vs. 5.3 +/- 2.2 mIU/mL; p=0.006), LH/FSH ratio (1.37 +/- 0.76 vs. 0.76 +/- 0.32; p<0.001), estrogen (54.5 +/- 18.2 vs. 38.0 +/- 11.1 pg/mL; p<0.001), testosterone (49.7 +/- 25.1 vs. 34.5 +/- 14.0 ng/dL; p=0.001), fasting insulin (11.55 +/- 10.8 vs. 5.63 +/- 2.6 uIU/mL; p=0.004), HOMA-IR (2.47 +/- 2.4 vs. 1.17 +/- 0.54; p=0.007), and leptin (31.3 +/- 17.4 vs. 16.4 +/- 10.5 ng/mL; p<0.001), with significantly lower FSH (5.6 +/- 1.7 vs. 7.2 +/- 1.9 mIU/mL; p<0.001) and HDL (43.0 +/- 10.0 vs. 54.3 +/- 15.3 mg/dL; p<0.001). Correlation analysis revealed positive associations between BMI and leptin, insulin, and HOMA-IR; WHR and testosterone; TLC and leptin/WHR; and LH/FSH ratio and estrogen, while HDL correlated negatively with HOMA-IR and TLC. Logistic regression identified leptin (OR = 1.105, 95% CI 1.016-1.201, p=0.020) and LH/FSH ratio (OR = 18.48, 95% CI 1.82-187.7, p=0.014) as independent predictors of PCOS.

**CONCLUSION:** Normoglycemic PCOS women show distinct hormonal, metabolic, and adipokine alterations, with leptin and LH/FSH emerging as robust independent predictors. These findings highlight the early convergence of adiposity, insulin resistance, inflammation, and gonadotropin imbalance in PCOS, underscoring the need for early biomarker-based risk stratification and intervention even before the onset of overt glycemic abnormalities.

## INTRODUCTION

Polycystic ovary syndrome (PCOS) is a common endocrinopathy affecting 5-20% of reproductive-aged women worldwide and is characterized by hyperandrogenism, ovulatory dysfunction, and polycystic ovarian morphology **(1, 2)**. Despite its heterogeneous clinical presentation, metabolic derangements including insulin resistance and adiposity-related abnormalities are recognized as central components of PCOS pathogenesis **(3, 4)**. Insulin resistance in PCOS contributes not only to glucose homeostasis disturbances but also exacerbates ovarian androgen production, thereby creating a vicious cycle of metabolic and reproductive dysfunction **(2, 5)**.

In addition to classical metabolic impairments, emerging evidence highlights the critical role of adipokines such as leptin, which mediate the cross-talk between adipose tissue, metabolism, and the hypothalamic-pituitary-ovarian (HPO) axis. Elevated leptin levels have been implicated in reproductive abnormalities and insulin resistance observed in PCOS, acting as a potential biomarker of disease severity **(6, 7)**. Moreover, dyslipidemia, characterized by reduced high-density lipoprotein (HDL) and adverse lipid profiles, is prevalent in PCOS and contributes to increased cardiovascular risk **(8)**.

Although insulin resistance and hyperandrogenism have been extensively studied in PCOS, there is limited data integrating hormonal, metabolic, inflammatory, and adipokine profiles specifically in normoglycemic women. Identifying early biomarkers and understanding their interplay is crucial for timely intervention to prevent progression to overt metabolic diseases.

Therefore, we conducted a comprehensive case-control study to evaluate the hormonal, metabolic, and adipokine profiles in normoglycemic PCOS women, and to assess the inter-relationships among adiposity, insulin resistance, inflammation, and gonadotropin imbalance. This study aims to elucidate early pathophysiological mechanisms in PCOS and identify potential independent predictors of the syndrome.

## OBJECTIVES

The primary objective of this study was to assess and compare the hormonal, metabolic, lipid, inflammatory and adipokine profiles between normoglycemic women diagnosed with PCOS and healthy controls. A secondary objective was to analyze the inter-relationships among adiposity, insulin resistance, gonadotropin imbalance, and adipokine levels in women with PCOS. Finally, we aimed to identify independent predictors of PCOS using multivariate logistic regression analysis.

## METHODS

### Study Design and Participants

This case-control study was conducted at Lady Hardinge Medical College between November, 2012 and April, 2014. Thirty women newly diagnosed with PCOS according to the Rotterdam criteria (2003) and 30 age-matched healthy normoglycemic controls were recruited. Inclusion criteria for cases included women aged 18-45 years with normoglycemia confirmed by fasting blood glucose and HbA1c within normal ranges. Controls were healthy women with regular menstrual cycles and no clinical or biochemical evidence of hyperandrogenism or insulin resistance.

### Ethical Approval

The study protocol was approved by the Institutional Ethics Committee of Lady Hardinge Medical College (LHMC), New Delhi and all participants provided written informed consent prior to enrolment.

### Clinical and Anthropometric Assessment

Detailed histories and physical examinations were performed. Anthropometric measurements including height, weight, body mass index (BMI), waist circumference, hip circumference, and waist-to-hip ratio (WHR) were recorded according to standardized protocols.

### Biochemical and Hormonal Assays

Fasting venous blood samples were collected on day 2-5 of menstrual cycle for the measurement of glucose, insulin, lipid profile (total cholesterol, triglycerides, HDL), HbA1c, complete blood count including total leukocyte count (TLC), and serum leptin. Hormonal assays included luteinizing hormone (LH), follicle-stimulating hormone (FSH), prolactin, estrogen, progesterone, and total testosterone. Insulin resistance was estimated using Homeostatic Model Assessment for Insulin Resistance (HOMA-IR) as HOMA-IR = [Fasting Glucose (mg/dL) × Insulin (µIU/mL)] / 405.

### Statistical Analysis

Data distributions were assessed using the Shapiro-Wilk test. Group comparisons between PCOS and controls were performed using independent samples t-test or Mann-Whitney U test as appropriate. Correlations between variables were analyzed using Pearson’s or Spearman’s correlation coefficients. Multivariate logistic regression was used to identify independent predictors of PCOS, including variables with significant univariate associations. Statistical significance was set at p<0.05. Analyses were conducted using SPSS version 30.

## RESULTS

### Baseline characteristics and group comparisons

Anthropometric indices of 30 normoglycemic women with PCOS when compared with 30 age-matched (mean age ∼25 years in both the groups) healthy controls revealed that women with PCOS had significantly higher body mass index (BMI) (26.3 ± 5.3 vs. 23.4 ± 3.2 kg/m^2^; p = 0.014) and waist-to-hip ratio (0.861 ± 0.061 vs. 0.799 ± 0.060; p < 0.001) compared with controls, indicating greater overall and central adiposity. (Table.1)

**Table.1:** Comparison of Anthropometric, Hormonal, Metabolic, Lipid, Inflammatory and Adipokine Markers between Normoglycemic Cases and Controls.

| Parameter | 30 Case (Mean ± SD) | 30 Control (Mean ± SD) | p-value |
| --- | --- | --- | --- |
| **BMI** | **$26.3 \pm 5.31$** | **$23.4 \pm 3.24$** | **0.014\*** |
| **WHR** | **$0.861 \pm 0.061$** | **$0.798 \pm 0.059$** | **<0.001\*\*** |
| TLC | $6856 \pm 1375$ | $6690 \pm 1668$ | 0.674 |
| **LH** | **$7.5 \pm 3.71$** | **$5.3 \pm 2.16$** | **0.006\*** |
| **FSH** | **$5.6 \pm 1.69$** | **$7.2 \pm 1.90$** | **<0.001\*\*** |
| **LH/FSH ratio** | **$1.37 \pm 0.76$** | **$0.76 \pm 0.32$** | **<0.001\*\*** |
| Prolactin | $18.21 \pm 9.17$ | $15.35 \pm 8.99$ | 0.112 |
| **Estrogen** | **$54.5 \pm 18.23$** | **$38.0 \pm 11.12$** | **<0.001\*\*** |
| Progesterone | $0.41 \pm 0.16$ | $0.46 \pm 0.21$ | 0.312 |
| **Testosterone** | **$49.7 \pm 25.1$** | **$34.5 \pm 14.04$** | **0.001\*** |
| Cholesterol | $166.4 \pm 42.21$ | $166.8 \pm 31.90$ | 0.455 |
| Triglyceride | $85.5 \pm 29.79$ | $89.9 \pm 29.45$ | 0.367 |
| **HDL** | **$43.0 \pm 10.01$** | **$54.3 \pm 15.25$** | **<0.001\*\*** |
| HbA1c | $4.69 \pm 0.82$ | $4.53 \pm 0.45$ | 0.554 |
| **Insulin** | **$11.55 \pm 10.81$** | **$5.63 \pm 2.56$** | **0.004\*** |
| **HOMA-IR** | **$2.47 \pm 2.40$** | **$1.17 \pm 0.54$** | **0.007\*** |
| **Leptin** | **$31.3 \pm 17.36$** | **$16.4 \pm 10.53$** | **<0.001\*\*** |

\*p-Value < 0.05 was considered significant. \*\*p-Value < 0.001 was considered highly significant.

Reproductive hormone analysis showed elevated luteinizing hormone (LH) (7.5 ± 3.7 vs. 5.3 ± 2.2 mIU/mL; p = 0.006) and a higher LH/FSH ratio (1.37 ± 0.76 vs. 0.76 ± 0.32; p < 0.001) in PCOS cases, along with lower follicle-stimulating hormone (FSH) (5.6 ± 1.7 vs. 7.2 ± 1.9 mIU/mL; p < 0.001). Serum estrogen and testosterone were significantly elevated in PCOS women (estrogen: 54.5 ± 18.2 vs. 38.0 ± 11.1 pg/mL, p < 0.001; testosterone: 49.7 ± 25.1 vs. 34.5 ± 14.0 ng/dL, p = 0.001), while progesterone and prolactin did not differ significantly.

Metabolic markers showed comparable fasting glucose (FBS) and HbA1c levels between groups; however, fasting insulin (11.55 ± 10.8 vs. 5.63 ± 2.6 µIU/mL; p = 0.004) and HOMA-IR (2.47 ± 2.4 vs. 1.17 ± 0.54; p = 0.007) were significantly higher in PCOS, indicating underlying insulin resistance. Lipid profile analysis revealed no significant differences in total cholesterol or triglycerides, but HDL was significantly lower in cases (43.0 ± 10.0 vs. 54.3 ± 15.3 mg/dL; p < 0.001).

Adipokine analysis demonstrated markedly higher leptin concentrations in women with PCOS compared to controls (31.3 ± 17.4 vs. 16.4 ± 10.5 ng/mL; p < 0.001). Total leukocyte count (TLC) and other basic biochemical parameters were comparable between groups.

### Correlation analysis within PCOS group

Correlation analysis revealed multiple significant associations highlighting interconnected endocrine-metabolic dysregulation:

- **BMI** correlated positively with leptin (r = 0.398, p = 0.029), insulin (ρ = 0.455, p = 0.012), and HOMA-IR (ρ = 0.427, p = 0.019).
- **WHR** correlated positively with testosterone (ρ = 0.427, p = 0.018).
- **TLC** showed a positive correlation with WHR (r = 0.429, p = 0.018) and leptin (r = 0.433, p = 0.017), while correlating negatively with HDL (ρ = −0.392, p = 0.032).
- **HDL** was inversely correlated with HOMA-IR (ρ = −0.365, p = 0.047).
- **Leptin** was significantly associated with HOMA-IR (ρ = 0.372, p = 0.043) and insulin (ρ = 0.410, p = 0.024).
- **LH/FSH ratio** correlated positively with estrogen (ρ = 0.423, p = 0.020).

### Multivariate Logistic Regression Analysis

A multivariate logistic regression model was performed including anthropometric, metabolic, hormonal, and adipokine variables as covariates (Table.3). Among the predictors, serum leptin (B = 0.100, OR = 1.105, 95% CI = 1.016-1.201, p = 0.020) and LH/FSH ratio (B = 2.917, OR = 18.48, 95% CI = 1.82-187.72, p = 0.014) emerged as significant independent predictors of PCOS. Neither BMI, WHR, insulin, HOMA-IR, nor testosterone remained significant after adjustment.

**Table.2:** Correlation between Anthropometric, Hormonal, Metabolic, Lipid, Inflammatory and Adipokine Markers within PCOS cases.

| Correlated Variables | Correlation coefficient | p-value | Direction |
| --- | --- | --- | --- |
| BMI-Leptin | 0.398 (r) | **0.029\*** | Positive |
| BMI-Insulin | 0.455 ($\rho$) | **0.012\*** | Positive |
| BMI-HOMA-IR | 0.427 ($\rho$) | **0.019\*** | Positive |
| WHR-Testosterone | 0.427 ($\rho$) | **0.018\*** | Positive |
| TLC-WHR | 0.429 (r) | **0.018\*** | Positive |
| TLC-Leptin | 0.433 (r) | **0.017\*** | Positive |
| TLC-HDL | -0.392 ($\rho$) | **0.032\*** | Negative |
| HDL-HOMA-IR | -0.365 ($\rho$) | **0.047\*** | Negative |
| Leptin-HOMA-IR | 0.372 ($\rho$) | **0.043\*** | Positive |
| Leptin- Insulin | 0.410 ($\rho$) | **0.024\*** | Positive |
| LH/FSH-Estrogen | 0.423 ($\rho$) | **0.020\*** | Positive |

\*p-value <0.05 was considered significant.

**Table.3:** Multivariate Logistic Regression Analysis for Predictors of PCOS.

| Predictor Variable | B (Coefficient) | SE | Wald $\chi^2$ | p-value | Odds Ratio (Exp(B)) | 95% CI for Exp(B) |
| --- | --- | --- | --- | --- | --- | --- |
| BMI | -0.061 | 0.106 | 0.328 | 0.567 | 0.941 | 0.764 - 1.159 |
| WHR | 9.378 | 8.094 | 1.343 | 0.247 | 11823.325 | 0.002 - 9.161 |
| **Leptin** | **0.100** | **0.043** | **5.443** | **0.020** | **1.105** | **1.016 - 1.201** |
| Insulin | 0.382 | 0.599 | 0.406 | 0.524 | 1.465 | 0.453 - 4.738 |
| HOMA-IR | -1.103 | 2.386 | 0.214 | 0.644 | 0.332 | 0.003 - 35.654 |
| **LH/FSH** | **2.917** | **1.183** | **6.081** | **0.014** | **18.480** | **1.819 - 187.719** |
| Testosterone | 0.035 | 0.022 | 2.494 | 0.114 | 1.036 | 0.992 - 1.082 |

\*p-value <0.05 was considered significant.

This indicates that for every unit increase in leptin, the odds of having PCOS increased by ∼10%, while women with higher LH/FSH ratios were nearly 18 times more likely to have PCOS independent of insulin resistance and adiposity.

### Subgroup Analysis: Lean vs. Overweight/Obese PCOS

Within the PCOS cohort, participants were stratified into lean (BMI <25 kg/m^2^) and overweight/obese (BMI ≥25 kg/m^2^) subgroups. Overweight/obese PCOS women exhibited significantly elevated fasting insulin (p=0.012), HOMA-IR (p=0.018), and leptin (p=0.007) compared to their lean counterparts, indicating greater insulin resistance and adipokine dysregulation linked to adiposity. (Table.4) However, reproductive hormone levels—including LH, FSH, LH/FSH ratio, and testosterone—did not differ significantly between lean and overweight/obese PCOS women, suggesting that reproductive endocrine dysfunction may manifest independently of BMI. No significant differences in lipid profile parameters (cholesterol, triglycerides, HDL) or leukocyte counts between lean and overweight/obese PCOS.

**Table.4:** Subgroup Analysis: Lean vs. Overweight/Obese PCOS (only significant parameters)

| Parameter | BMI < 25 (Mean ± SD) | BMI ≥ 25 (Mean ± SD) | p-value |
| --- | --- | --- | --- |
| WHR | 0.8337 ± 0.0643 | 0.8865 ± 0.0484 | 0.047* |
| TLC | 6250 ± 1199.8 | 7387.63 ± 1329.78 | 0.022* |
| Insulin | 7.35 ± 4.73 | 15.23 ± 13.25 | 0.031* |
| HOMA-IR | 1.54 ± 1.10 | 3.27 ± 2.93 | 0.028* |
| Leptin | 24.91 ± 18.00 | 36.94 ± 15.16 | 0.031* |

\*p-value <0.05 was considered significant.

### BMI-Matched Analysis: PCOS vs. Controls in BMI ≥25 Subgroup

Comparing PCOS cases and BMI-matched controls within the overweight/obese subgroup revealed significantly higher waist-to-hip ratio (0.889 ± 0.049 vs. 0.846 ± 0.052; p=0.047), LH (6.76 ± 2.76 vs. 4.51 ± 1.55; p=0.029), estrogen (53.09 ± 13.78 vs. 37.43 ± 10.72; p=0.006), and testosterone (48.77 ± 16.38 vs. 28.53 ± 14.16; p=0.004) in PCOS cases. FSH was significantly lower (5.66 ± 1.07 vs. 7.57 ± 2.02; p=0.005), resulting in a higher LH/FSH ratio (1.22 ± 0.57 vs. 0.62 ± 0.24; p=0.005). Markers of insulin resistance, including fasting insulin (15.97 ± 13.37 vs. 5.67 ± 3.56; p=0.027) and HOMA-IR (3.43 ± 2.97 vs. 1.21 ± 0.79; p=0.031), were also elevated in PCOS cases. Additionally, leptin levels were significantly higher in the overweight/obese PCOS group compared to controls (37.64 ± 15.43 vs. 22.60 ± 15.11; p=0.024) (Table.5)

**Table.5:** BMI-Matched Analysis: PCOS vs. Controls in BMI ≥25 Subgroup (only significant parameters)

| Parameter | PCOS Cases (BMI $\geq 25$)<br>(Mean ± SD) | Controls (BMI $\geq 25$)<br>(Mean ± SD) | p-value |
| --- | --- | --- | --- |
| WHR | $0.889 \pm 0.049$ | $0.846 \pm 0.052$ | 0.047* |
| LH | $6.76 \pm 2.76$ | $4.51 \pm 1.55$ | 0.029* |
| FSH | $5.66 \pm 1.07$ | $7.57 \pm 2.02$ | 0.005* |
| LH/FSH | $1.22 \pm 0.57$ | $0.62 \pm 0.24$ | 0.005* |
| Estrogen | $53.09 \pm 13.78$ | $37.43 \pm 10.72$ | 0.006* |
| Testosterone | $48.77 \pm 16.38$ | $28.53 \pm 14.16$ | 0.004* |
| Insulin | $15.97 \pm 13.37$ | $5.67 \pm 3.56$ | 0.027* |
| HOMA-IR | $3.43 \pm 2.97$ | $1.21 \pm 0.79$ | 0.031* |
| Leptin | $37.64 \pm 15.43$ | $22.60 \pm 15.11$ | 0.024* |

\*p-value $<0.05$ was considered significant.

## DISCUSSION

### Reproductive Hormonal Findings

In our study, women with PCOS demonstrated significantly elevated LH and LH/FSH ratio, with concomitantly lower FSH, compared to controls. This gonadotropin imbalance is a hallmark of PCOS, reflecting altered hypothalamic-pituitary-ovarian axis dynamics that perpetuate anovulation and infertility. Elevated estrogen and testosterone further confirmed ovarian steroidogenic dysfunction, in line with earlier reports of hyperandrogenism and estrogen excess as consistent biochemical features of PCOS **(1, 9, 10)**. The positive correlation between LH/FSH and estrogen strengthens the hypothesis that gonadotropin dysregulation promotes estrogenic overactivity, contributing to abnormal folliculogenesis **(11, 12)**.

### Metabolic Disturbances

Although fasting glucose and HbA1c were comparable between groups, fasting insulin and HOMA-IR were significantly higher in the PCOS group. This indicates that insulin resistance precedes hyperglycemia in PCOS, a finding consistently reported in diverse ethnic populations **(3, 4, 13)**. Insulin resistance plays a central role in exacerbating androgen production by theca cells and reducing sex-hormone binding globulin, thereby worsening hyperandrogenemia **(14, 15)**.

Lipid analysis revealed significantly lower HDL among PCOS women, despite similar total cholesterol and triglyceride levels. Reduced HDL is an early feature of atherogenic dyslipidemia in PCOS and contributes to heightened cardiovascular risk **(8, 15)**. Although our cohort was normoglycemic and relatively young (mean age ∼25 years), this lipid alteration highlights how metabolic risk emerges early in the disease trajectory.

### Adipokines and Inflammation

Leptin concentrations were markedly increased in PCOS women and correlated significantly with BMI, insulin, HOMA-IR, and TLC. These associations underscore leptin’s role as an adiposity-linked endocrine signal that bridges obesity and reproductive function. Hyperleptinemia and leptin resistance in PCOS may impair normal folliculogenesis and ovulation **(16, 17)**. Additionally, leptin correlated with TLC, supporting its involvement in sustaining chronic low-grade inflammation — an important pathophysiological feature of PCOS linked to both reproductive and cardiometabolic outcomes **(18, 19)**.

The observed negative association between HDL and both TLC and HOMA-IR further reinforces the interplay between inflammation, insulin resistance, and dyslipidemia, consistent with reports of immune-metabolic crosstalk in PCOS **(10, 20)**.

### Multivariate logistic regression analysis

The regression model further underscored the central role of leptin and LH/FSH ratio in distinguishing PCOS from controls. Elevated leptin independently predicted PCOS, even after controlling for BMI and insulin resistance, suggesting that leptin dysregulation contributes to the pathophysiology of PCOS beyond simple adiposity. This is consistent with studies indicating that leptin resistance may disrupt neuroendocrine regulation and directly impair ovarian function **(21, 22)**.

The LH/FSH ratio retained strong predictive power, highlighting its enduring diagnostic and pathogenic relevance. An OR of 18.5 underscores the clinical utility of this simple biochemical marker as an independent discriminator of PCOS, in agreement with earlier reports of HPO axis dysregulation as a cornerstone feature **(1)**.

Interestingly, traditional metabolic indicators such as insulin, HOMA-IR, BMI, and WHR lost significance in the multivariate model, reflecting that while these are altered in PCOS, their predictive capacity diminishes when leptin and LH/FSH are taken into account. This finding suggests that a combination of gonadotropin imbalance and adipokine signaling may be the most robust predictors of PCOS in normoglycemic women **(7, 23)**.

### BMI-Stratified Analysis

The present study’s stratified analyses elucidate important aspects of the heterogeneity inherent in PCOS and its relationship with adiposity. The comparison between lean and overweight/obese PCOS women demonstrates that metabolic disturbances—specifically hyperinsulinemia, elevated HOMA-IR, and increased leptin—are more pronounced with higher adiposity. This is in line with established evidence that obesity exacerbates insulin resistance and adipokine dysregulation in PCOS **(3, 24)**. Conversely, reproductive hormone abnormalities including elevated LH, altered LH/FSH ratios, and hyperandrogenism appear largely unaffected by BMI status, corroborating findings from previous studies suggesting that endocrine dysfunction in PCOS is intrinsic and not solely a consequence of adiposity **(1, 25)**.

In the BMI-matched case–control analysis within the overweight/obese subgroup, PCOS women continued to exhibit significantly elevated LH, increased LH/FSH ratio, hyperandrogenism, insulin resistance, and leptin compared to controls despite similar BMI levels. This supports the notion that the metabolic and hormonal perturbations characteristic of PCOS are not merely secondary to obesity but represent inherent pathophysiological features **(2, 4, 25)**. The modestly higher waist-to-hip ratio observed in the overweight/obese PCOS group aligns with literature emphasizing the role of central adiposity as a contributor to metabolic risk beyond total body weight **(8, 26)**.

Leptin’s significant elevation and its persistence after BMI adjustment reinforce its emerging role not only as an adiposity marker but also as a mediator of reproductive and metabolic dysfunction in PCOS. Elevated leptin has been linked with impaired neuroendocrine regulation of ovulation and may contribute to insulin resistance through inflammatory pathways **(6, 7, 10)**. Similarly, the robust elevation of LH/FSH ratio underscores continued hypothalamic– pituitary–ovarian axis dysregulation, a hallmark of PCOS central to follicular arrest and anovulation **(1, 27)**.

## CLINICAL IMPLICATIONS

The findings of this study have important clinical implications for the diagnosis and management of PCOS, particularly in normoglycemic women. Recognition that key reproductive hormonal abnormalities and metabolic disturbances persist irrespective of BMI underscores the necessity to evaluate all PCOS patients comprehensively, not only those who are visibly overweight or obese **(1, 28)**. Importantly, measuring fasting insulin levels—even in women with normal glucose tolerance—is critical for early detection of insulin resistance, which often precedes overt hyperglycemia and plays a central role in PCOS pathophysiology **(3, 4, 29)**.

Early assessment of leptin levels alongside traditional hormonal markers such as the LH/FSH ratio and fasting insulin can improve risk stratification, allowing clinicians to identify women at higher metabolic and cardiovascular risk despite normal glycemic status **(8, 11, 30)**. Personalized treatment strategies should therefore address both metabolic and endocrine dysfunctions, with lifestyle modification and insulin-sensitizing therapies considered essential regardless of BMI or glycemic measurements **(31)**. Additionally, monitoring adipokines such as leptin may provide further insights into metabolic health and treatment response **(32)**. Integrating these biomarkers into routine clinical practice can enhance early intervention, ultimately improving reproductive and metabolic outcomes for women with PCOS.

## CONCLUSION

This study highlights that normoglycemic women with PCOS exhibit significant metabolic and hormonal abnormalities regardless of their BMI status. While adiposity amplifies insulin resistance and leptin levels, key reproductive hormonal dysfunctions such as elevated LH/FSH ratio and hyperandrogenism are intrinsic features independent of obesity. The persistence of these abnormalities in BMI-matched comparisons with controls underscores the complex, multisystem nature of PCOS. Leptin and LH/FSH ratio emerge as robust, obesity-independent markers that may aid early diagnosis and personalized management. Tailored interventions addressing both metabolic and reproductive disturbances are essential for improving outcomes across the PCOS spectrum.

## LIMITATIONS

The limitations of this study are a relatively small sample size (30 cases and 30 controls), which may limit statistical power and generalizability. The cross-sectional design prevents causal inferences regarding adipokine and hormonal changes in PCOS. We did not assess additional factors such as inflammatory cytokines, detailed body composition, or genetic profiles, which could further clarify underlying mechanisms. Lastly, recruitment from a single tertiary centre may introduce selection bias toward more severe cases, limiting external validity. Future larger, multicentre, longitudinal studies incorporating broader biomarkers are needed to validate and extend these findings.

## Data Availability

All data produced in the present study are available upon reasonable request to the authors

## ACKNOWLEDGMENTS

The Authors would like to thank Dr Chitra Raghunandan, Former Director Professor, Dept of Obst and Gynae, LHMC, for providing cases for the study. The authors also wish to thank the faculty, senior residents, junior residents and technical & other staff at Department of Biochemistry, LHMC, New Delhi for their assistance and support in data collection and analysis.

## References

1. Azziz R CE, Chen Z, Dunaif A, Laven JSE, Legro RS, et al. Polycystic ovary syndrome. Nat Rev Dis Primers. 2016;2:16057.

2. Sanchez-Garrido MA T-SM. Metabolic dysfunction in polycystic ovary syndrome: Pathogenic role of androgen excess and potential therapeutic strategies. Molecular Metabolism. 2020;35:100937.

3. A.D. Insulin resistance and the polycystic ovary syndrome: mechanism and implications for pathogenesis. Endocr Rev. 1997;18(6):774–800.

4. Moran LJ MM, Wild RA, Norman RJ. Impaired glucose tolerance, type 2 diabetes and metabolic syndrome in polycystic ovary syndrome: a systematic review and meta-analysis. Hum Reprod Update. 2010;16(4):347–63.

5. Chen W PY. Metabolic Syndrome and PCOS: Pathogenesis and the Role of Metabolites. Metabolites. 2021;11(12):869.

6. Nikanfar S OH, Rastgar Rezaei Y, Zarezadeh R, Jafari-gharabaghlou D, Nejabati HR, et al. Role of adipokines in the ovarian function: Oogenesis and steroidogenesis. The Journal of Steroid Biochemistry and Molecular Biology. 2021;209(105852).

7. Schüler-Toprak S OO, Buechler C, Treeck O. The Complex Roles of Adipokines in Polycystic Ovary Syndrome and Endometriosis. Biomedicines. 2022;10:2503.

8. RA. W. Dyslipidemia in PCOS. Steroids. 2012;77(4):295–9.

9. Miller WL AR. The molecular biology, biochemistry, and physiology of human steroidogenesis and its disorders. Endocr Rev. 2011;32(1):81–151.

10. Magoffin DA. Totowa N. Ovarian steroidogenic abnormalities in the polycystic ovary syndrome. Totowa, NJ.: Humana Press.; 2006 2006 Sep 1.

11. Longo M LF, De Carlini S, La Marca A.;23(Suppl 1):22. The role of LH in follicle development: from physiology to new clinical implications. Reprod Biol Endocrinol. 2025;23:22.

12. Hajam YA RH, Kumar R, Basheer M, Reshi MS.. A review on critical appraisal and pathogenesis of polycystic ovarian syndrome. Endocrine and Metabolic Science. 2024;14:100162.

13. Dong J RD. Polycystic ovary syndrome: pathophysiology and therapeutic opportunities. BMJ medicine. 2023;2(1):e000548.

14. Ding H ZJ, Zhang F, Zhang S, Chen X, Liang W, Xie Q.. Resistance to the Insulin and Elevated Level of Androgen: A Major Cause of Polycystic Ovary Syndrome. Front Endocrinol (Lausanne). 2021 Oct 20;12:741764.

15. Unluhizarci K KZ, Kelestimur F.. Role of insulin and insulin resistance in androgen excess disorders. World J Diabetes. 2021;12(5):616–29.

16. Peng Y YH, Song J, Feng D, Na Z, Jiang H, Meng Y, Shi B, Li D.. Elevated serum leptin levels as a predictive marker for polycystic ovary syndrome. Frontiers in Endocrinology. 2022;13(845165).

17. J. C. Serum leptin level in women with polycystic ovary syndrome: correlation with adiposity, insulin, and circulating testosterone. Ann Med Health Sci Res. 2013;3(2):191–6.

18. Sneha S HS. The Role of Inflammatory Pathways in PCOS-Related Infertility and Pregnancy Complications. European Journal of Cardiovascular Medicine. 2024;14:679–84.

19. Pérez-Pérez A S-JF, Vilariño-García T, Sánchez-Margalet V.. Role of Leptin in Inflammation and Vice Versa. Int J Mol Sci. 2020;21(16):5887.

20. Asmathulla S RV, Kripa S, Rajarajeswari R.. Insulin resistance and its relation to inflammatory status and serum lipids among young women with PCOS. International Journal of Reproduction, Contraception, Obstetrics and Gynecology. 2013;2(3):325–30.

21. Chou SH MC. Role of leptin in human reproductive disorders. J Endocrinol. 2014;223(1):T49–62.

22. Farooqi IS ORS. 20 years of leptin: human disorders of leptin action. Journal of Endocrinology. 2014;223(1):T63–70.

23. Merza WM YA, Mahmood MA.. FSH, LH, lipid and adipokines in polycystic ovary syndrome: clinical biochemistry insights for diagnosis and management. The Journal of Steroid Biochemistry and Molecular Biology. 2025:106773.

24. Kumari M KS, Das J.. Adipokine Dysregulation in Obese and Non-Obese Polycystic Ovary Syndrome (PCOS) Patients: Association With Visceral Adiposity Index and Metabolic Risk. Cureus. 2025;17(7).

25. Barber TM HP, Weickert MO, Franks S.. Obesity and Polycystic Ovary Syndrome: Implications for Pathogenesis and Novel Management Strategies. Clin Med Insights Reprod Health. 2019;13:1179558119874042.

26. Rashmi S HN, Reddy V, Veena BM. Evaluation of body composition in body mass index matched PCOS and eumenorrheic non-PCOS college women. International Journal of Community Medicine and Public Health. 2025;12(4):1682.

27. Liao B QJ, Pang Y. Central regulation of PCOS: abnormal neuronal-reproductive-metabolic circuits in PCOS pathophysiology. Frontiers in endocrinology. 2021;12:667422.

28. Zhang H WW, Zhao J, Jiao P, Zeng L, Zhang H, Zhao Y, Shi L, Hu H, Luo L, Fukuzawa I Relationship between body composition, insulin resistance, and hormonal profiles in women with polycystic ovary syndrome. Frontiers in Endocrinology. 2023;13:1085656.

29. Diamanti-Kandarakis E DA. Insulin resistance and the polycystic ovary syndrome revisited: an update on mechanisms and implications. Endocrine reviews. 2012;33(6):981–1030.

30. Singhal AK SG, Singh SK, Karunanand B, Gunjan G, Agrawal SK. Exploring the link between leptin levels and metabolic syndrome in elderly Indian patients: Implications for family medicine and primary care practices. J Family Med Prim Care. 2024;13(9):3633–8.

31. Kalra S JB, Yeravdekar R. Emotional and Psychological Needs of People with Diabetes. Indian J Endocrinol Metab. 2018;22(5):696–704.

32. M B. Adipokines–removing road blocks to obesity and diabetes therapy. Molecular metabolism. 2014;3(3):230–40.

